# Climate-Sensitive Health Outcomes in Kenya: A Scoping Review of Environmental Exposures and Health Outcomes Research, 2000 – 2023

**DOI:** 10.1101/2024.04.25.24306268

**Authors:** Jessica Gerard, Titus Kibaara, Iris Martine Blom, Jane Falconer, Shamsudeen Mohammed, Zaharat Kadri-Alabi, Roz Taylor, Leila Abdullahi, Robert C Hughes, Bernard Onyango, Ariel A Brunn

## Abstract

Climate change threatens health and social development gains in Kenya, necessitating health policy planning for risk reduction and mitigation. To understand the baseline state of knowledge of environmental determinants of health in Kenya relevant to climate change, a comprehensive scoping review was undertaken. Compliant with a pre-registered protocol, nine bibliographic databases and grey literature sources were searched for articles published from 2000-2023. Two-stage screening was conducted on 17,394 articles; 635 full-texts were screened in duplicate. A final 353 articles underwent data extraction for topic categorisation, bibliometric analysis, and narrative summary. This was comprised of 344 (97%) journal articles, 59% of which were published after 2014 (n=207). Main study designs included observational (n=211) and modelling studies (n=64). Health topics centred on vector-borne diseases (41%, n=147), primarily vector abundance (n=102) and malaria (n=60), while injury or death (n=10), mental health conditions (n=7) and heat exposure (n=7) studies were less frequent. Environmental health research in Kenya is largely conducted in the Lake Victoria Basin, Rift Valley and Coastal regions, with fewer studies from the northern arid and semi-arid regions. Findings of this review suggest a growing and diverse field of predominantly observational research with an increasing focus on social determinants and policy-relevant themes, however research on vector-borne disease dwarfs other health outcomes and sparsely populated but climatically fragile regions are less represented in published literature. Addressing existing gaps in baseline evidence underpinning associations between the environment and health outcomes will benefit climate change attribution research and support future development of evidence-informed climate change and health policy in Kenya.

## Introduction

The recent convergence of increasing extreme weather events and rising awareness of attributable anthropogenic contributions to climate change has sharpened global attention to the impact of environment on human health, wellbeing and livelihoods. This is particularly evident in sub-Saharan Africa (SSA) which experiences adverse effects of climate change, despite nominal contributions to global greenhouse gas emissions [1]. Kenya is highly susceptible to climate change due to its varied topography, diverse climatic zones, and reliance on natural resources and is the largest economy in East Africa with a growing population of 2.3% projected to nearly double by the turn of the century [2].

Currently, Kenya stands at an inflection point toward rapid industrialisation and sustainable growth, with the potential to provide a blueprint for climate-resilient economic development in the region. Kenya was the first country in Africa to enact legislation exclusively on climate change via the 2016 Climate Change Act [3], which sets out pathways towards sustainable development through the National Climate Change Action Plan. This plan in turn advises on mechanisms of integrating sectoral climate change mitigation and adaptation actions at national and sub-national levels [4]. In 2023, Kenya convened the inaugural Africa Climate Summit in conjunction with the Africa Union Commission and launched the Nairobi declaration on green development [5] which linked health alongside economic development in support of the United Nations 2030 Sustainable Development Goals [6].

Various environmental exposures (EE) including weather, hydrometeorological hazards and air pollution pose risks to social development gains due to their influence on human health [7]. However, many of these causal relationships have not been clearly defined in Kenya, potentially limiting opportunities for detection and attribution research over longer timeframes that would permit the evaluation of health impacts of climate change, with implications for the development of evidence-informed climate change and health policy. To better understand the state of environmental health research, a comprehensive synthesis of published output on the influence of EE on health outcomes (HO) in Kenya was undertaken with the following objectives: (a) to undertake a scoping review of literature on relationships between EE and HO; (b) to map the links between these exposures and climate-sensitive HO and health equity through bibliometric analysis, topic mapping and narrative synthesis; and (c) to identify knowledge gaps and future research needs to strengthen the evidence base underpinning climate change and health (CCH) attribution for policy development. This article presents initial bibliometric analysis and narrative summary findings from the broader scoping review work on environmental health research in Kenya.

## Materials and Methods

### Protocol and registration

Reporting of this review was guided by the PRISMA extension for scoping reviews [8], as well as an established team of article reviewers, a library information professional, and subject matter experts in medicine, veterinary medicine, nutrition, demography, water, sanitation and health, and child health and development. A scoping review was deemed most suitable given the complexities and wide breadth of the subject matter and lack of similar reviews [9]. The protocol for this review was registered on Open Science Framework on April 14, 2023 [10].

### Eligibility criteria

We aimed to identify a wide range of original literature describing the relationships between EE and HO, excluding intervention studies. The range of exposures included weather variables such as temperature and precipitation, hydrometeorological hazards including droughts and flooding, and climate variability phenomena such as El Niño – Southern Oscillation, with a full list found in Table S1. In recognition of the moderating effects of land-use change, terrestrial, aquatic and air pollution on health, these environmental drivers were also incorporated. Eligible health outcomes encompassed direct and indirectly impacted outcomes for example, heat stroke and vector-borne diseases (VBD), respectively.

We included any original research published in English between January 1^st^, 2000, and February 20, 2023. This timeframe reflects the growing interest and discourse on health implications of climate change as well as improvements in environmental attribution methods in public health sciences [11]. As our review focused on Kenya, we included studies on any demographic populations including transborder pastoralist communities, as well as global studies, if data from Kenya was disaggregated and extractable. Eligible studies were required to include some measure of a HO produced by either qualitative or quantitative analysis.

### Information source

The search strategy was informed by the Population-Exposure-Comparator-Outcome (PECO) model: [12]

**P:** population of Kenya

**E:** environmental exposures, including weather, hydrometeorological hazards and air quality variables

**C:** no effect of environmental exposures on health conditions (as available, studies will not be excluded for lack of comparison groups)

**O:** disease burdens or measures of association or effect of environmental exposures on health outcomes and included synonyms for health outcomes guided by categories listed in the World Health Organisation report, Quality Criteria for Health National Adaptation Planning [13]. The full methods, search terms and database search results were conducted by a Library information professional and are hosted in an open access digital repository maintained by the London School of Hygiene & Tropical Medicine [14]. Nine bibliographic databases were searched in February 2023: Medline, Embase, Global Health, Food Science and Technology Abstract and Econlit via OvidSP, GreenFile and Africa-Wide Information via EBSCOhost, Clarivate Analytics Web of Science core content and Scopus. Grey literature sources included Google Scholar and websites of 10 organizations known to be working on environment or CCH research in Kenya.

### Selection process

All citations were deduplicated and transferred into the reference manager software EPPI-Reviewer Web 4.14.2.0 [15] for two-stage screening. Title and abstract screening was conducted by a single trained reviewer against *a priori* inclusion criteria. All reviewers were trained on an initial sample of 800 abstracts in duplicate to ensure consistency. Following primary screening, full-text articles were reviewed in duplicate for eligibility. At both stages of screening, reasons for exclusion were noted.

### Data extraction

Data extraction was undertaken by two independent reviewers and confirmation of final extracted data arbitrated by a third reviewer. All articles were categorized by main HO (Table 1) and EE (Table S1). Main categories were further divided into subcategories that were refined using an iterative approach during data extraction. Data on publication year; author institutional affiliation; study type; article type; funder(s); location(s); and analysis method(s) were recorded. To investigate authorship and geographical extent of collaborations, the institutional affiliation(s) of authors were categorized into regional or international groups; funders were likewise allocated into one of six funding models. Analytical methods for each article were assessed by main study design and analysis methods employed. Covariate results were extracted and described in brief narrative summaries.

**Table 1.** Health outcome topic categories and their corresponding subcategories.

| Topic Category | Topic Subcategories |  |
| --- | --- | --- |
| Injury or Death | Direct injury<br>Direct death | Burden estimate |
| Heat Exposure & Skin Conditions | Heat stress<br>Skin conditions |  |
| Cardiovascular, Circulatory & Respiratory Disorders | Heart disease & circulatory disorders<br>Lung & airway conditions | Respiratory infections |
| Waterborne Diseases & Water Access Disorders | Cholera<br>Diarrheal diseases<br>Leptospirosis<br>Cryptosporidiosis<br>Schistosomiasis | Giardia<br>Dehydration & kidney disorders<br>Water insecurity<br>Harmful algal blooms |
| Vector-borne Diseases | Vector or parasite abundance or prevalence<br>Malaria<br>Dengue<br>Trypanosomiasis<br>Lymphatic filariasis<br>Leishmaniasis | Tick-borne diseases<br>Soil-transmitted helminths<br>Yellow Fever<br>Chikungunya<br>West Nile Virus |
| Zoonoses | Animal reservoir abundance or zoonotic disease prevalence<br>Anthrax<br>*Coxiella burnetti* | Bartonellosis<br>Rift Valley fever<br>Brucellosis |
| Malnutrition & Foodborne Diseases | Stunting<br>Wasting<br>Malnutrition | Food insecurity<br>*Escherichia coli* & *Salmonella* |
| Mental Health Conditions | Eco-anxiety & depression<br>Stress & resilience | Cognitive capacity |
| Adverse Birth or Pregnancy Outcomes | Neonatal or infant outcomes<br>Maternal health outcomes |  |
| Health Equity Research | Toxic level: air pollutants<br>Toxic level: water & terrestrial pollutants<br>Toxic level: other pollutants<br>Neoplasia<br>Pre-existing conditions<br>Displacement & migration<br>Occupational hazards | Health vulnerability<br>Awareness & Perceptions<br>Conflict<br>Gender-based violence<br>Access to health facilities<br>Health inequities |

### Evidence synthesis

We used bibliometric analysis and narrative summaries to synthesize the evidence. To explore characteristics of environment and health research in Kenya, we mapped locations of empirical research on HO to Kenya’s main climatological zones [16], used keywords for conceptual mapping using VOSviewer software [17] and developed a Sankey diagram to illustrate environmental drivers of HO using R software [18, 19].

### Protocol amendments

We applied two protocol amendments to our review. We expanded our eligibility criteria to include a category of research articles that measured pollutant toxicity since they provided data on an exposure risk, even if they did not measure impact on an explicit HO.

The second protocol amendment was made to the HO categories informed by the Quality Criteria for Health National Adaptation Plans [13]. We added the category “Adverse Pregnancy or Birth Conditions” to better reflect gender disparities in HO and expanded equity related subthemes under a category titled “Health Equity”.

## Results

The bibliographic database search identified 29,443 records, of which 12,132 were duplicates removed prior to screening (Fig 1). The grey literature search identified 77 pieces of grey literature and 6 records from cited references, resulting in 17,394 unique references that underwent title and abstract screening. Of these, 635 underwent full text screening and 353 met inclusion criteria for the final article set (Table S2). A total of 26 full text reports, including 13 conference abstracts, could not be retrieved.

**Fig 1.**
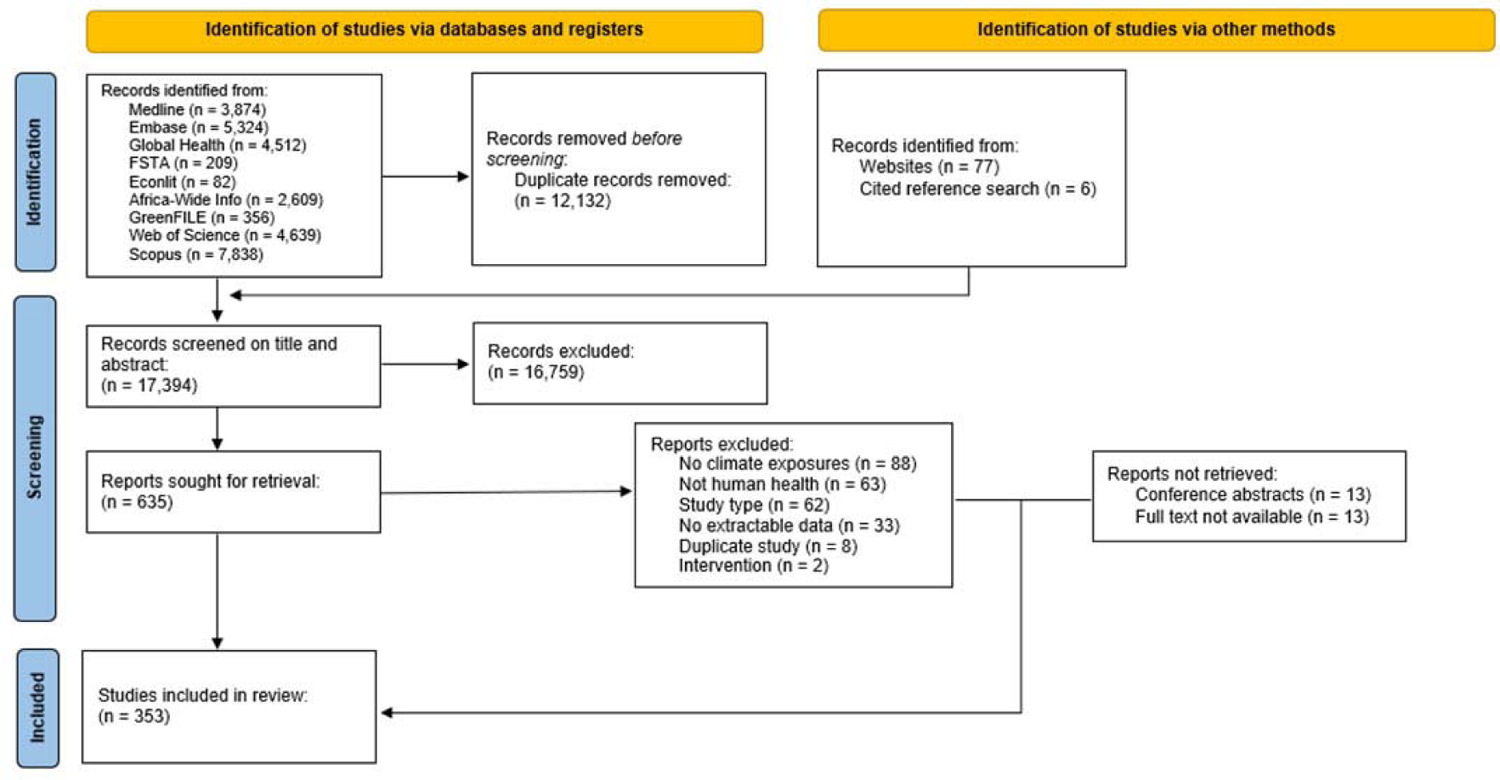
PRISMA flow diagram of evidence selection [20].

### Article characteristics

The characteristics of the final set of articles are described in Table 2. The study set is primarily comprised of journal articles (97%, n=344). Research on environmental determinants of health in Kenya increased in frequency per 5-year period from 2000 to 2019 with a small decline between 2020–2023, likely due to the shorter search period of 3 years and 2 months. Analysis of author’s institutional affiliations found that 52% (n=186) of articles were authored by international collaborations, while fewer than 20% (n=67) were exclusively from Kenyan institutions. Likewise, the majority of funding came from international public and private research funders, with just over 10% (n=46) of funders cited as public (n=28), university (n=10), or private funders (n=8) from Kenya. 58 studies did not cite a funding source.

**Table 2.** Single selectable bibliometric characteristics of included literature.

| Characteristic | Included literature (n) |
| --- | --- |
| **Year Published** | **353** |
| 2000 - 2004 | 25 |
| 2005 - 2009 | 43 |
| 2010 - 2014 | 78 |
| 2015 - 2019 | 118 |
| 2020 - 2023 | 89 |
| **Institutional Collaborations** | **353** |
| Kenyan | 67 |
| African Region | 26 |
| International | 186 |
| International without African collaborations | 74 |
| **Study Type** | **353** |
| Observational | 211 |
| Modelling Study | 64 |
| Mixed methods | 24 |
| Trials* | 24 |
| Qualitative | 27 |
| Meta-analysis | 3 |
| **Geographic Scale** | **353** |
| National | 53 |
| Regional | 194 |
| City/community | 91 |
| Not specified/relevant | 13 |

\*Includes both randomised and non-randomised trials

Most study designs (60%, n=211) were observational in nature while modelling, qualitative, and randomized and non-randomized trials constituted 18% (n=64), 7.6% (n=27), and 6.7% (n=24), respectively. Methods of analysis reflected the main study designs, where 21% (n = 144/689, Table 3) used regression analysis, 8% (n=54/689) advanced modelling methods including mechanistic modelling, spatial modelling, machine learning and multicriteria decision making, 6.5% (n = 45/689) qualitative methods such as interviews and focus groups and 2.1% (n = 15/689) applied health risk exposure assessment calculations.

**Table 3.**
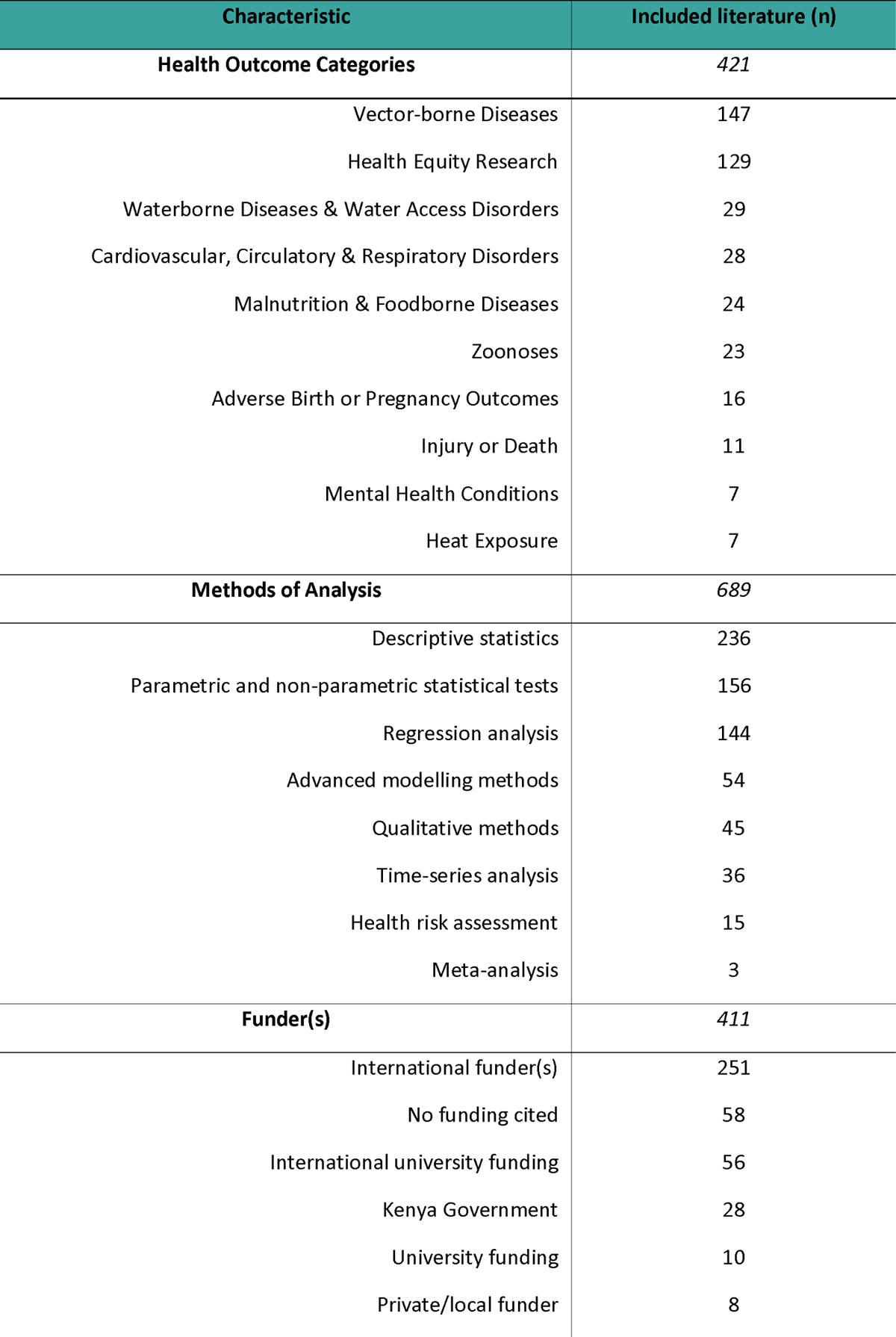
Multi-selectable bibliometric characteristics of included literature.

| Characteristic | Included literature (n) |
| --- | --- |
| **Health Outcome Categories** | **421** |
| Vector-borne Diseases | 147 |
| Health Equity Research | 129 |
| Waterborne Diseases & Water Access Disorders | 29 |
| Cardiovascular, Circulatory & Respiratory Disorders | 28 |
| Malnutrition & Foodborne Diseases | 24 |
| Zoonoses | 23 |
| Adverse Birth or Pregnancy Outcomes | 16 |
| Injury or Death | 11 |
| Mental Health Conditions | 7 |
| Heat Exposure | 7 |
| **Methods of Analysis** | **689** |
| Descriptive statistics | 236 |
| Parametric and non-parametric statistical tests | 156 |
| Regression analysis | 144 |
| Advanced modelling methods | 54 |
| Qualitative methods | 45 |
| Time-series analysis | 36 |
| Health risk assessment | 15 |
| Meta-analysis | 3 |
| **Funder(s)** | **411** |
| International funder(s) | 251 |
| No funding cited | 58 |
| International university funding | 56 |
| Kenya Government | 28 |
| University funding | 10 |
| Private/local funder | 8 |

Table 2 summarises publication year, institutional collaboration, study type, and geographic scale. Articles were allocated once within each category.

Table 3 summarises health outcome categories, methods of analysis and funder type. Articles could be allocated more than once within each category.

### Keyword trends

To identify trends in research interest, bibliometric keyword co-occurrence analysis was used to map the temporal relationship between terms in publication titles and abstracts [17]. A total of 11,070 terms were identified and for optimization purposes, a threshold of inclusion of a minimum of 8 occurrences was established and a relevance score of 60% was used, resulting in 142 keywords. The period of time over which the greatest shift in keyword trends occurred is shown in Fig 2. Connection lines indicate networks and circle sizes correspond to occurrence. Cluster density visualization highlights closeness between terms over time, illustrated as two clusters of keywords: cluster 1 (left, n = 68) VBD and climate exposures and cluster 2 (right, n = 74) social science and policy-oriented keywords.

**Fig 2.**
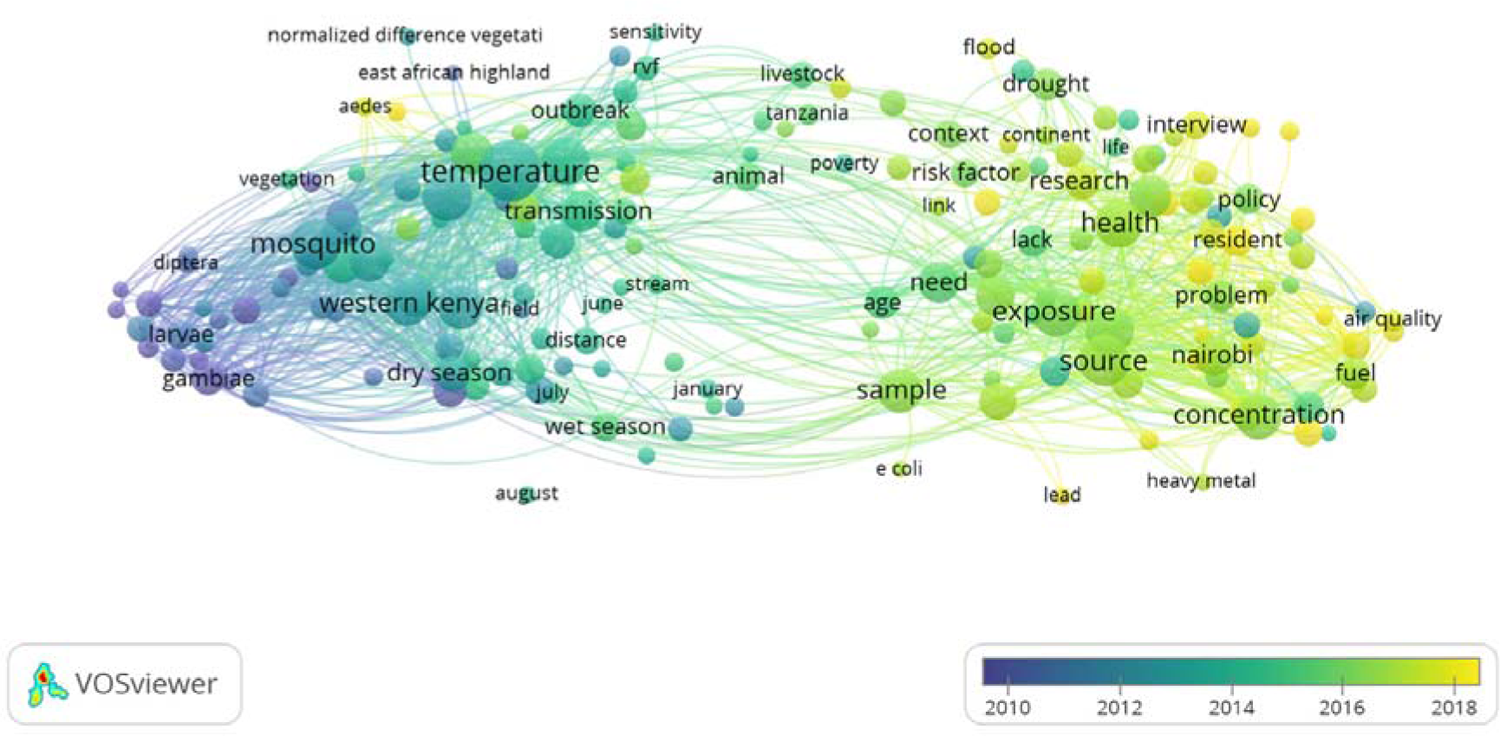
Time-scaled bibliometric keyword analysis.

A network map based on co-occurrence of keywords in titles & abstracts of included articles.

### Environmental drivers of health outcomes in Kenya

Occurrences of environmental covariates of HO are shown in Fig 3, extracted from 353 articles. Rainfall represented the most frequent exposure studied (n=218) and was specifically investigated as a driver of VBD in 115 occurrences within the article set. Other exposures linked to VBD included temperature (n=118), habitat change (n=61) and seasonality (n=60) – the latter of which was the third-most studied exposure (n=142) after temperature (n=168). Climate change was a non-specific exposure term used in studies that evaluated awareness of participants to climate change impacts on health (n=10). Less studied EE by frequency included wildfires (n=1), plastic pollution (n=1) and water level change (n=3).

**Fig 3.**
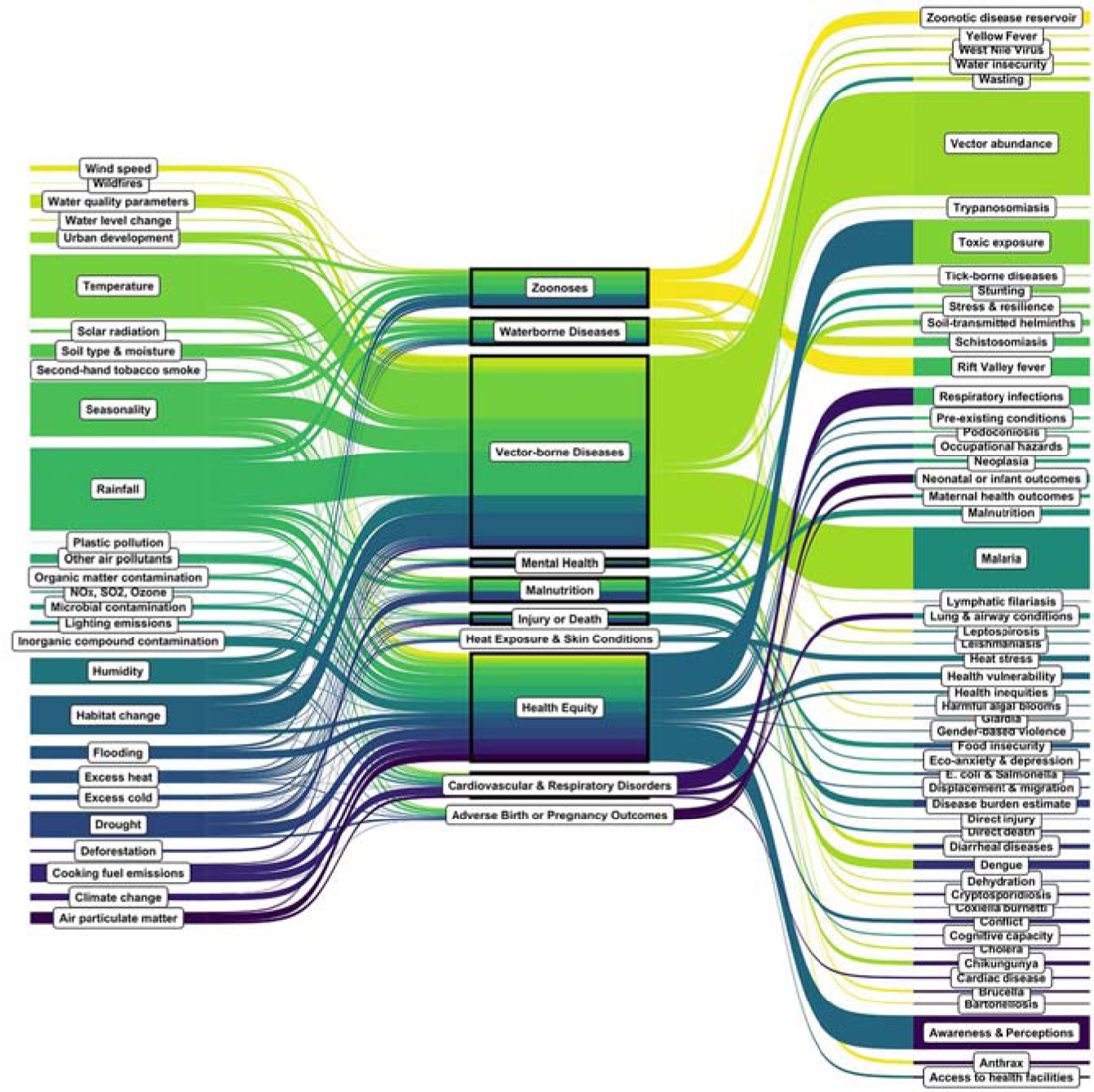
Sankey diagram of pathways between environmental exposures and health outcomes. Some category and subcategory names have been abbreviated for illustrative purposes, see table 1 for full categorization.

The most frequently studied HO was VBD (n=502), followed by research relevant to health equity (n=281). The health equity category encompassed research describing toxic levels of air, terrestrial and water pollutants (n=117) (see Table 1 for full listing). Health outcomes including heat stress (n=15), maternal health outcomes (n=9), access to health facilities (n=6), eco-anxiety and depression (n=4) were infrequent in the article set. A full listing of all studies categorised by main HO is provided in Table S2.

### Research locations in Kenya

Health outcome topic mapping from 287 articles with sub-location data (Fig 4) highlights the geographical distribution of empirical research based on climatological zones in Kenya [16]. The highest density of research occurs in Kenya’s humid southwest region, including the Lake Victoria Basin, Rift Valley region and Nairobi, where 75% of VBD (153/205) and 80% of health equity research (105/132) was conducted. In contrast, HO studied in the northern arid and semi-arid lands (ASALs) of Kenya more frequently focused on malnutrition and zoonoses, of which less literature was identified overall (Table 3). Empirical research was least frequently conducted in the North Western Region, where it was also the least diverse of all regions, encompassing only half of all possible outcome categories.

**Fig 4.**
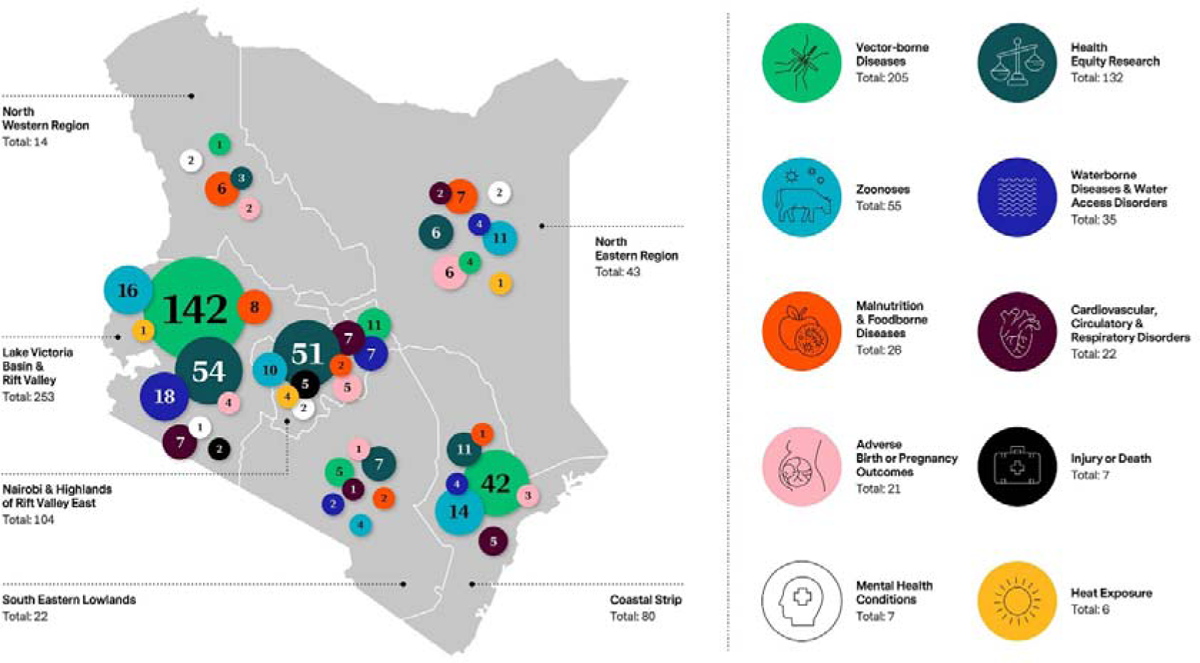
Health outcomes studied in empirical environmental research conducted in Kenya by climatological zone.

This figure shows health outcome results extracted from studies that cited a location for data collection (n=287).

### Health outcome summaries

#### Vector-borne diseases

VBDs were the most studied HO in this review, evaluated in 147 studies, 69% of which focused on vector abundance (n=102). These studies included assessments of malaria, dengue, yellow fever, West Nile Virus and other VBD, and explored a variety of burden indicators, such as disease risk, incidence, and vector population dynamics influenced by EE. Malaria was the most studied disease, based on both clinical case reports (n=60) and vector abundance studies (n=66), with most research conducted in southwestern Kenya (Table S2). There was comparatively less literature available on non-malarial diseases such as dengue (n=7); soil-transmitted helminths (n=4); West Nile Virus (n=2) and a single result each for trypanosomiasis, lymphatic filariasis, leishmaniasis, tick-borne diseases and yellow fever.

#### Health equity research

The largest subtopic of this category assessed respondent’s awareness and perceptions of CCH (n=51), followed by exposure to air and water pollution, with 36 and 22 articles respectively. Demographic and social determinants were frequent elements to these studies, especially air pollutant exposure studies in informal settlements [21, 22] and studies on gender vulnerabilities of indoor air pollution [23–25]. Gendered access or lack thereof was identified in educational attainment and accessibility to health facilities [26, 27]. Two studies explored gender roles and violence in view of climate change, identifying a potential exacerbation of inequalities and harmful practices like female genital mutilation and intimate partner violence [27, 28]. Water-scarcity driven conflicts in pastoralist communities were also explored in three studies [29–31].

#### Waterborne diseases & water access disorders

Twenty-nine studies investigated waterborne disease and water access disorders in Kenya. Warming temperatures both increased and decreased Schistosomiasis transmission depending on the region, while precipitation had delayed influence on snail vector density [32–34]. Increased rainfall during the rainy season impacted cholera risk [35], and seasonality significantly influenced snail abundance and cryptosporidium prevalence [36–38]. Three studies on diarrheal disease risk assessed in the under-five population reported that dry season trends drive rotavirus infections [39–41].

#### Cardiovascular, circulatory & respiratory disorders

A number of studies on cardiovascular, circulatory and respiratory disorders assessed the relationship with household air pollutants (HAPs) (n=14), finding that exposure, especially over extended periods of time in women and children is linked to adverse outcomes [23, 42]. Exposure to HAPs such as particulate matter (PM_2.5_) and carbon monoxide (CO) have been linked to reduced cardiac function [43] and volatile organic compounds from wood smoke are associated with increased self-reported respiratory, eye irritation and headache symptoms [44]. In households that used firewood or unprocessed biomass, children and infants under the age of five had a greater relative risk of developing acute respiratory infections (ARI) compared to those using kerosene fuels; long-term exposure to PM_2.5_ also increased these conditions and symptoms [42, 45, 46].

#### Malnutrition & foodborne diseases

Approximately 5% of studies included in this review evaluated the impact of climate variables on nutritional deficiencies (n=24), frequently measured in relation to drought or changes in precipitation or through proxy measures such as Normalized Difference Vegetation Index. In northern Kenyan counties, temperature is positively associated with malnutrition [47] and the impact of drought is inversely correlated with validated growth measures such as middle-upper-arm-circumference, height-for-age and weight-for-age Z-scores in children [48, 49].

#### Zoonoses

Two studies assessed how EE impact wildlife host density and immunocompetence of rodents in relation to zoonotic disease [50], as well as interactions between precipitation and land-use change on infected rodent host density [51]. Climate risk factors for Rift Valley Fever (RVF), a zoonotic virus that causes disease in livestock and haemorrhagic fever in humans, was evaluated in fifteen studies through empirical and modelling methods applied to livestock, humans and mosquito vectors. Rainfall abnormalities, vegetation change and measures of humidity were shown to determine the abundance and suitability of potential habitats for RVF vectors [52–54], and also impact vector and host susceptibility [55].

#### Adverse birth or pregnancy outcomes

Environmental exposures that impact the health of mothers and infants were investigated in 16 studies. Seven articles evaluated precipitation, seasonality and drought and the associated effect on anthropometric measurements of nutritional status [49, 56]. Disease prevalence in children under five years of age, assessed in six studies, was characterized by seasonal patterns of occurrence; investigated diseases included rotavirus, *Escherichia coli,* shigellosis, cryptosporidiosis and other enteropathogens [37, 39–41]. Two additional studies looked at infant ARI, including human metapneumovirus [57] and Respiratory Syncytial Virus incidence [58].

#### Injury or death

Ten studies were categorized as evaluating death or injury due to trauma linked to environmental exposures as well as studies that measured disease burden, using standard mortality and morbidity metrics. The relationship between temperature and premature mortality was assessed using years of life lost [59, 60] and through burden estimates of morbidity [61–63] and disability-adjusted life-years (DALYs) [64]. DALYs were also used in four studies to estimate the overall burden of HAPs on health [64–67].

#### Mental health conditions

Mental health conditions were infrequently studied (n = 7) in the article set. These studies evaluated the impacts of extreme weather events and climate shocks on economic status, mental wellbeing, and psychological distress [68–71].

#### Heat exposure & skin conditions

Four studies investigated the impacts of temperature extremes on mortality measures and found significant positive associations between exposure to low temperature and mortality in Nairobi populations [59, 60, 72, 73]. An additional two studies reported on vulnerability indices [74], hydration in agro pastoralists [75] and a single study described Podoconiosis distribution and risk prediction in relation to different scenarios of environmental suitability [76].

## Discussion

Our review compiled literature on EE and health impacts in Kenya, and in doing so, identified broad trends and emerging gaps in evidence. With meteorological projections indicating warmer temperatures, changing rainfall patterns and increasing flood and drought events [77, 78], the findings of this review are relevant to researchers and policymakers aiming to establish the risks to population health attributable to Kenya’s changing climate.

### Research trends

We found a diverse range of research on environmentally mediated HO. Nonetheless, the highest proportion of literature focused on VBD, particularly studies that evaluated clinical malaria as well as malaria vector abundance. There is an identified inequality in global funding trends for research on the 23 WHO-defined neglected tropical diseases (NTDs), as emphasized by funding from United States of $100 million on these diseases versus $1.5 billion for human immunodeficiency virus, malaria and tuberculosis combined in 2016 alone [79–81]. While this funding has substantially advanced understanding and supported successful control measures of these high burden diseases, many environmentally mediated NTDs in Kenya remain understudied [82]. In addition, a comparatively low amount of literature used a One Health approach to evaluate interconnected relationships between human, animal and environmental health pathways. This may be due to a lack of data, difficulties in establishing ecological dependencies, and challenges in the use of integrative and multisectoral approaches to support sustainable control efforts [83].

Most literature in this review was published by collaborations of international authors and supported by international funders, raising questions about inequities in global health research and funding structures in Kenya particularly given the prominence of Kenyan actors in CCH research in Africa [84]. More equitable funding and support for Kenyan-produced research outputs could drive contextually relevant research agendas and better amplify local voices [84, 85].

### Shifting narratives

Our evaluation of key themes in environment and health research in Kenya suggests a broadening of interest over the last decade towards social science,health equity and policy research from more conventional environmental health topics such as VBD. This shift reflects a larger pattern of change towards improvements in attribution methods alongside cross-disciplinary approaches, including those in social sciences, occurring in public health research [86–88]. Subjects relevant to health equity were identified in several articles focused on vulnerable populations including pollution exposure studies of residents of informal settlements, water scarcity conflicts in pastoralist communities, and extreme weather effects on intimate partner violence, cognitive development and education access for young girls. Some studies measured associations between EE and HO, such as air pollution exposure on cardiovascular or respiratory outcomes, but a proportion measured toxic exposure risks only. A shift was also seen towards studies which explored contextual experience of CCH pathways, assessed through qualitative techniques. Perceptions of how climate change impacts health provide insights to the need for improved public health promotion in Kenya and underscore calls for targeted capacity building as well as climate change training for healthcare workers and the wider public health sector [89].

### Research gaps

Mental health conditions were less frequently studied in our article set. A recent scoping review found that despite rapid growth, the global output of climate change and mental health research is comparatively lower than other health conditions and limited in scope [90]. Similarly, heat exposure was understudied, despite climate model projections that indicate parts of Kenya, alongside other SSA countries, will experience the greatest increase in frequency of heat stress days globally [74].

Recent research from West Africa has confirmed a link between heat impacts and adverse birth outcomes related to fetal strain [91], however most research on this topic is based on data from high-income countries, possibly due to scarcity of temperature ground monitoring and health data [92]. Given the high fertility rate in Kenya and relatively high rate of neonatal mortality [93, 94], the low volume of gender-oriented articles points to an important research gap. Accordingly, we found need to amend health outcome categories that were based on a WHO framework on climate-sensitive health risks to specifically include adverse birth and pregnancy outcomes [95], given recognized inequities in climate impacts on women’s health [96].

There was a gap in research on malnutrition and foodborne disease, a principal risk factor of deaths and disabilities in Kenya, despite evidence that climate change has contributed to the ongoing Horn of Africa drought causing 20,000 excess child deaths in 2022 [97–99]. In infants and children, malnutrition can have long-lasting health impacts which is relevant to Kenya’s young population [48]. In the northern ASALs of Kenya where pastoralist communities reside, there is a need for greater exploration of the impacts of worsening drought patterns on these groups [100]. These EE can cause secondary effects in communities in the form of tribal conflict over water and livestock resources or gendered violence [27, 31]. Ongoing conflict and insecurity in northern areas may exacerbate geographic disparities seen in research output, resulting in a much higher density of publications in the southwestern region, home to nearly 90% of the population in an area less than 20% of Kenyan land mass [2, 101]

#### Limitations

As environmental health research is a rapidly growing area of study, our search terminology attempted to encompass a wide variety of exposures and outcomes in line with WHO defined pathways. We aimed to minimize missing articles by using broad terminology in our search and screening criteria, incorporating a grey literature search, and using recognized frameworks for health categorization. In addition, this scoping review did not set out to conduct a risk of bias assessment and thus cannot draw conclusions on quality of evidence; however future reviews would benefit from assessment to provide confidence in results.

## Conclusions

This review provides a baseline analysis of the scale and scope of evidence describing environmental impacts on health in Kenya. Its diversity illustrates the wide range of health pathways that have been studied in Kenya and identifies trends in institutional collaborations, funding patterns, and research priorities. Greater attention is needed on vulnerable groups, geographical disparities in research and complex relationships between environmental determinants and less frequently studied HO to ensure equity of the growing research. Targeted capacity building, funding reform and enhanced support for local and regional institutional networks are necessary steps to build the evidence base and safeguard population health in the face of Kenya’s changing climate.

### CRediT authorship contribution statement

**Jessica Gerard**: Formal Analysis, Investigation, Visualization, Data Curation, Writing – original draft, Writing – Review & Editing. **Titus Kibaara**: Investigation, Data Curation, Writing – original draft, Writing – Review & Editing. **Iris Blom:** Conceptualization, Methodology, Investigation, Data Curation, Writing – Review and Editing. **Jane Falconer:** Methodology, Investigation, Data Curation, Writing – Review & Editing. **Shamsudeen Mohammed:** Methodology, Investigation, Data Curation, Writing – Review & Editing. **Zaharat Kadri-Alabi**: Investigation, Data Curation, Writing – Review & Editing. **Roz Taylor:** Investigation, Data Curation, Writing – Review & Editing. **Leila Abdullahi:** Conceptualization, Methodology, Writing – Review & Editing. **Robert C Hughes:** Conceptualization, Investigation, Writing – Reviewing & Editing. **Bernard Onyango**: Conceptualization, Investigation, Writing – Reviewing & Editing. **Ariel Brunn**: Conceptualization, Formal Analysis, Investigation, Visualization, Writing – original draft, Writing – Review & Editing, Supervision.

### Declaration of competing Interest

IMB declares having received three partial grants for her studies. A Prince Bernhard Culture Fund grant number 40037327 was awarded on 15 September 2021; Stichting VSBFonds grant number VSB.21/00168 was awarded on 17 May 2021; dr. Hendrik Mullerfonds without grant number was awarded on 9 December 2021. RCH reports receiving grant funding for research on climate change and health from the Wellcome Trust, The Bernard van Leer Foundation, and Fondation Botnar, and has worked as an employee or consultant for the Children’s Investment Fund Foundation, The Clean Air Fund and The Abu Dhabi Early Childhood Development Authority. Additional authors declare they have no actual or potential competing interests that could influence the work herein reported.

## Supporting information

Supplemental Table 1 of environmental exposure topic categories and their corresponding subcategories

Supplemental Table 2 of included study set narrative summaries

PRISMA-ScR Checklist

## Data Availability

The full methods, search terms and database search results are hosted in an open access digital repository maintained by the London School of Hygiene & Tropical Medicine. Citation: Falconer, J and Brunn, A (2024). Search strategies for "Scoping study of Economic and Data System Considerations for Climate Change and Pandemic Preparedness in Africa". [Data Collection]. London School of Hygiene & Tropical Medicine, London, United Kingdom. https://doi.org/10.17037/DATA.00003769.

## Acknowledgements

This work was supported by the Childrens’ Investment Fund Foundation [grant number 2008-05023]. The funders had no role in study design, data collection and analysis, decision to publish, or preparation of this manuscript. The opinions expressed here belong to the authors, and do not necessarily reflect those of the funder.

## Supporting Information

**S1 Table.**
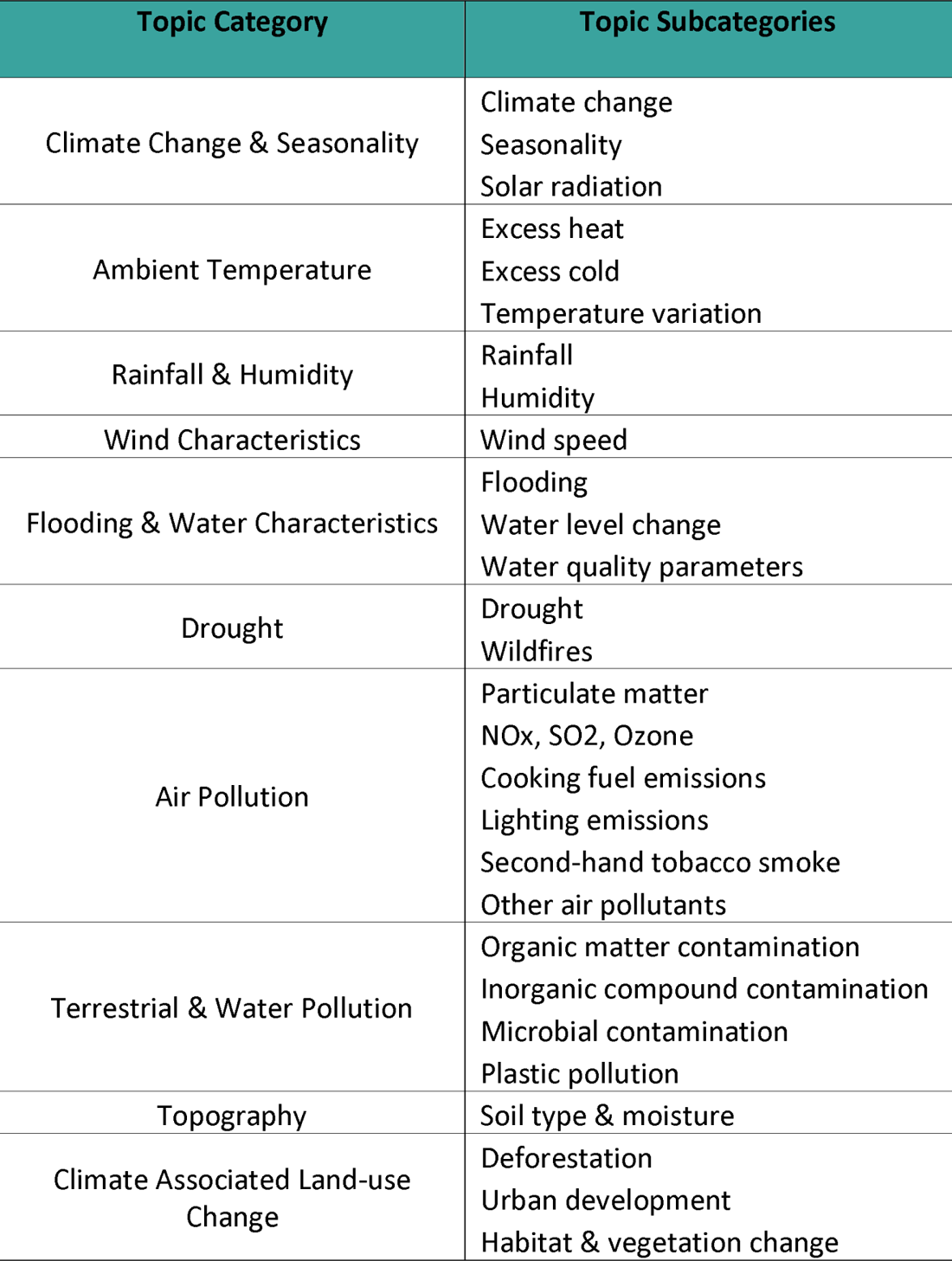
Environmental exposure topic categories and their corresponding subcategories.

**S2 Table.** Narrative summaries table Included study set with summaries of results and information pertinent to this research.

## Notes

### Clinical Protocols

https://osf.io/fp6sd

## References

1. World Meteorological Organization. State of the Climate in Africa 2021. 2022. Available from: https://library.wmo.int/records/item/58070-state-of-the-climate-in-africa-2021.

2. World Bank Group. Climate Risk Profile: Kenya. 2021. Available from: https://climateknowledgeportal.worldbank.org/sites/default/files/2021-05/15724-WB_Kenya%20Country%20Profile-WEB.pdf.

3. Wambua C. The Kenya Climate Change Act 2016 Emerging Lessons From a Pioneer Law. Carbon & Climate Law Review. 2019;13(4):257–69, doi:10.21552/cclr/2019/4/6.

4. Government of the Republic of Kenya. National Climate Change Action Plan (Kenya): 2018-2022. In: Ministry of Environment and Forestry, editor. Nairobi2018. Available from: https://rise.esmap.org/data/files/library/kenya/Clean%20Cooking/Kenya_NCCAP_2018-2022.pdf.

5. Oguta E. Kenya Hosts First African Climate Summit. Diplomacy Newsletter [Internet]. 2023; (25). Available from: https://mfa.go.ke/wp-content/uploads/2023/10/DIPLOMACY-ENEWSLETTER-AUG-SEP-EDITION.pdf.

6. United Nations. Transforming our World: The 2030 Agenda for Sustainable Development. 2015. Available from: https://sdgs.un.org/2030agenda.

7. Mora C, McKenzie T, Gaw IM, Dean JM, Von Hammerstein H, Knudson TA, et al. Over half of known human pathogenic diseases can be aggravated by climate change. Nature Climate Change. 2022;12(9):869–75, doi:10.1038/s41558-022-01426-1.

8. Sarkis-Onofre R, Catalá-López F, Aromataris E, Lockwood C. How to properly use the PRISMA Statement. Systematic Reviews. 2021;10(1):117, doi:10.1186/s13643-021-01671-z.

9. Pham MT, Rajić A, Greig JD, Sargeant JM, Papadopoulos A, McEwen SA. A scoping review of scoping reviews: advancing the approach and enhancing the consistency. Research Synthesis Methods. 2014;5(4):371–85, doi:10.1002/jrsm.1123.

10. Brunn A, Ray S, Blom I, Gerard J, Hughes R.C., Kibaara T., Onyango B, Abdullahi L, Falconer J. Climate-Sensitive Health Outcomes in Kenya: A Protocol for a Scoping Review of the Impact of Environmental Exposures on Health Outcomes. 2023, doi: 10.17605/OSF.IO/FP6SD.

11. Callaghan MW, Minx JC, Forster PM. A topography of climate change research. Nature Climate Change. 2020;10(2):118–23, doi:10.1038/s41558-019-0684-5.

12. Morgan RL, Whaley P, Thayer KA, Schünemann HJ. Identifying the PECO: A framework for formulating good questions to explore the association of environmental and other exposures with health outcomes. Environ Int. 2018;121(Pt 1):1027–31, doi:10.1016/j.envint.2018.07.015.

13. World Health Organization. Quality criteria for health national adaptation plans. Geneva 2021. Available from: https://www.who.int/publications/i/item/9789240018983.

14. Falconer J, Brunn A. Search strategies for “A Scoping Review of the Influence of Environmental Exposures on Health Outcomes in Kenya”. London, United Kingdom: London School of Hygiene & Tropical Medicine. 2023, doi:10.17037/DATA.00003705.

15. Thomas J, Brunton, J., Graziosi, S. EPPI-Reviewer 4.0: software for research synthesis. EPPI Centre Software. London: Social Science Research Unit, Institute of Education, University of London; 2010.

16. Kenya Meteorological Department. State of the Climate in Kenya 2020. [Internet]. 2020. Available from: www.meteo.go.ke.

17. van Eck NJW, L. VOSViewer: Visualizing Scientific Landscapes: Version 1.6.19. 2023.

18. R Core Team. R: A language and environment for statistical computing. R Foundation for Statistical Computing. Vienna, Austria2023.

19. Popay J, Roberts H, Sowden A, Petticrew M, Arai L, Rodgers M, Britten N, Roen K, Duffy S. Guidance on the conduct of narrative synthesis in systematic reviews. ESRC Methods Programme Version 12006. p. 1–92. Available from: https://www.lancaster.ac.uk/media/lancaster-university/content-assets/documents/fhm/dhr/chir/NSsynthesisguidanceVersion1-April2006.pdf.

20. Page MJ, McKenzie JE, Bossuyt PM, Boutron I, Hoffmann TC, Mulrow CD, et al. The PRISMA 2020 statement: an updated guideline for reporting systematic reviews. BMJ. 2021;372:n71, doi:10.1136/bmj.n71.

21. Egondi T, Muindi K, Kyobutungi C, Gatari M, Rocklöv J. Measuring exposure levels of inhalable airborne particles (PM2.5) in two socially deprived areas of Nairobi, Kenya. Environ Res. 2016;148:500–6, doi:10.1016/j.envres.2016.03.018.

22. Kiai C, Kanali C, Sang J, Gatari M. Spatial Extent and Distribution of Ambient Airborne Particulate Matter (PM(2.5)) in Selected Land Use Sites in Nairobi, Kenya. J Environ Public Health. 2021;2021:4258816, doi:10.1155/2021/4258816.

23. Dohoo C, Guernsey JR, Critchley K, VanLeeuwen J. Pilot study on the impact of biogas as a fuel source on respiratory health of women on rural Kenyan smallholder dairy farms. J Environ Public Health. 2012;2012:636298, doi:10.1155/2012/636298.

24. Ezzati M, Saleh H, Kammen DM. The contributions of emissions and spatial microenvironments to exposure to indoor air pollution from biomass combustion in Kenya. Environ Health Perspect. 2000;108(9):833–9, doi:10.1289/ehp.00108833.

25. Ochieng C, Vardoulakis S, Tonne C. Household air pollution following replacement of traditional open fire with an improved rocket type cookstove. Sci Total Environ. 2017;580:440–7, doi:10.1016/j.scitotenv.2016.10.233.

26. Zuurmond M, Nyapera V, Mwenda V, Kisia J, Rono H, Palmer J. Childhood disability in Turkana, Kenya: Understanding how carers cope in a complex humanitarian setting. Afr J Disabil. 2016;5(1):277, doi:10.4102/ajod.v5i1.277.

27. Esho T, Komba, E., Richard, F., Shell-Duncan, B. Intersections between climate change and female genital mutilation among the Maasai of Kajiado County, Kenya. Journal of Global Health. 2021(1), doi:10.7189/jogh.11.04033.

28. Allen EM, Munala L, Henderson JR. Kenyan Women Bearing the Cost of Climate Change. Int J Environ Res Public Health. 2021;18(23), doi:10.3390/ijerph182312697.

29. Linke AM, O’Loughlin J, McCabe JT, Tir J, Witmer FDW. Rainfall variability and violence in rural Kenya: Investigating the effects of drought and the role of local institutions with survey data. Global Environmental Change. 2015;34:35–47, doi:10.1016/j.gloenvcha.2015.04.007.

30. Linke AM, Witmer FDW, O’Loughlin J. Weather variability and conflict forecasts: Dynamic human-environment interactions in Kenya. Political Geography. 2022;92:102489, doi:10.1016/j.polgeo.2021.102489.

31. Omosa E. The Impact of Water Conflicts on Pastoral Livelihoods: The Case of Wajir District in Kenya.: International Institute for Sustainable Development; 2005. Available from: https://www.iisd.org/publications/report/impact-water-conflicts-pastoral-livelihoods-case-wajir-district-kenya.

32. McNally KL. Developing risk assessment maps for Schistosoma haematobium in Kenya based on climate grids and remotely sensed data: Louisiana State University; 2003. Available from: https://repository.lsu.edu/gradschool_theses/1851.

33. McCreesh N, Nikulin G, Booth M. Predicting the effects of climate change on Schistosoma mansoni transmission in eastern Africa. Parasites & Vectors. 2015;8(1):4, doi:10.1186/s13071-014-0617-0.

34. Kariuki HC, Clennon JA, Brady MS, Kitron U, Sturrock RF, Ouma JH, et al. Distribution patterns and cercarial shedding of Bulinus nasutus and other snails in the Msambweni area, Coast Province, Kenya. Am J Trop Med Hyg. 2004;70(4):449–56, doi: 10.4269/ajtmh.2004.70.449.

35. Stoltzfus JD, Carter JY, Akpinar-Elci M, Matu M, Kimotho V, Giganti MJ, et al. Interaction between climatic, environmental, and demographic factors on cholera outbreaks in Kenya. Infect Dis Poverty. 2014;3(1):37, doi:10.1186/2049-9957-3-37.

36. Muchiri JM, Ascolillo L, Mugambi M, Mutwiri T, Ward HD, Naumova EN, et al. Seasonality of Cryptosporidium oocyst detection in surface waters of Meru, Kenya as determined by two isolation methods followed by PCR. J Water Health. 2009;7(1):67–75, doi:10.2166/wh.2009.109.

37. Gatei W, Wamae CN, Mbae C, Waruru A, Mulinge E, Waithera T, et al. Cryptosporidiosis: prevalence, genotype analysis, and symptoms associated with infections in children in Kenya. Am J Trop Med Hyg. 2006;75(1):78–82, doi: 10.4269/ajtmh.2006.75.78.

38. Mutuku FM, King CH, Bustinduy AL, Mungai PL, Muchiri EM, Kitron U. Impact of drought on the spatial pattern of transmission of Schistosoma haematobium in coastal Kenya. Am J Trop Med Hyg. 2011;85(6):1065–70, doi:10.4269/ajtmh.2011.11-0186.

39. Omore R, Tate JE, O’Reilly CE, Ayers T, Williamson J, Moke F, et al. Epidemiology, Seasonality and Factors Associated with Rotavirus Infection among Children with Moderate-to-Severe Diarrhea in Rural Western Kenya, 2008-2012: The Global Enteric Multicenter Study (GEMS). PLoS One. 2016;11(8):e0160060, doi:10.1371/journal.pone.0160060.

40. Lambisia AW, Onchaga S, Murunga N, Lewa CS, Nyanjom SG, Agoti CN. Epidemiological Trends of Five Common Diarrhea-Associated Enteric Viruses Pre- and Post-Rotavirus Vaccine Introduction in Coastal Kenya. Pathogens. 2020;9(8), doi:10.3390/pathogens9080660.

41. Gikonyo J, Mbatia B, Okanya P, Obiero G, Sang C, Nyangao J. Rotavirus prevalence and seasonal distribution post vaccine introduction in Nairobi county Kenya. Pan Afr Med J. 2019;33:269, doi:10.11604/pamj.2019.33.269.18203.

42. Sikolia DN, Mwololo, K., Cherop, H., Hussein, A., Juma, M., Kurui, J., Bwika, A., Seki, I., Osaki, Y. The prevalence of acute respiratory infections and the associated risk factors: a study of children under five years of age in Kibera Lindi Village, Nairobi, Kenya. Journal of National Institute of Public Health. 2002;51(1):67–72. Available from: https://www.niph.go.jp/journal/data/51-1/200251010012.pdf.

43. Agarwal A, Kirwa K, Eliot MN, Alenezi F, Menya D, Mitter SS, et al. Household Air Pollution Is Associated with Altered Cardiac Function among Women in Kenya. Am J Respir Crit Care Med. 2018;197(7):958–61, doi:10.1164/rccm.201704-0832LE.

44. Critchley K. Air quality, respiratory health and wood use for women converting from low- to high-efficiency stoves in rural Kenya. 2015, doi:10.2495/air150171.

45. Foote EM, Gieraltowski L, Ayers T, Sadumah I, Faith SH, Silk BJ, et al. Impact of locally-produced, ceramic cookstoves on respiratory disease in children in rural western Kenya. Am J Trop Med Hyg. 2013;88(1):132–7, doi:10.4269/ajtmh.2012.12-0496.

46. Larson PS, Espira L, Glenn BE, Larson MC, Crowe CS, Jang S, O’Neill MS. Long-Term PM(2.5) Exposure Is Associated with Symptoms of Acute Respiratory Infections among Children under Five Years of Age in Kenya, 2014. Int J Environ Res Public Health. 2022;19(5), doi:10.3390/ijerph19052525.

47. Kipkulei H, Boitt M, Imwatis A. Spatial Variability of Malnutrition and Predictions Based on Climate Change and Other Causal Factors: A Case Study of North Rift ASAL Counties of Kenya. Journal of Earth Science & Climatic Change. 2017;8(10):416, doi:10.4172/2157-7617.1000416.

48. Bauer JM, Mburu S. Effects of drought on child health in Marsabit District, Northern Kenya. Econ Hum Biol. 2017;24:74–9, doi:10.1016/j.ehb.2016.10.010.

49. Ongudi ST, D R. Prenatal health and weather-related shocks under social safety net policy in Kenya. Economic Research Southern Africa (ERSA). 2020;831. Available from: https://econrsa.org/wp-content/uploads/2022/06/working_paper_831.pdf.

50. Young HS, McCauley DJ, Dirzo R, Nunn CL, Campana MG, Agwanda B, et al. Interacting effects of land use and climate on rodent-borne pathogens in central Kenya. Philosophical Transactions of the Royal Society B: Biological Sciences. 2017;372(1722):20160116, doi:10.1098/rstb.2016.0116.

51. Young HS, Dirzo R, Helgen KM, McCauley DJ, Billeter SA, Kosoy MY, et al. Declines in large wildlife increase landscape-level prevalence of rodent-borne disease in Africa. Proc Natl Acad Sci U S A. 2014;111(19):7036–41, doi:10.1073/pnas.1404958111.

52. Anyamba A, Chretien JP, Small J, Tucker CJ, Formenty PB, Richardson JH, et al. Prediction of a Rift Valley fever outbreak. Proc Natl Acad Sci U S A. 2009;106(3):955–9, doi:10.1073/pnas.0806490106.

53. Campbell LP, Reuman DC, Lutomiah J, Peterson AT, Linthicum KJ, Britch SC, et al. Predicting Abundances of Aedes mcintoshi, a primary Rift Valley fever virus mosquito vector. PLOS ONE. 2019;14(12):e0226617, doi:10.1371/journal.pone.0226617.

54. Mosomtai G, Evander M, Sandström P, Ahlm C, Sang R, Hassan OA, et al. Association of ecological factors with Rift Valley fever occurrence and mapping of risk zones in Kenya. Int J Infect Dis. 2016;46:49–55, doi:10.1016/j.ijid.2016.03.013.

55. McIntyre KM, Setzkorn C, Hepworth PJ, Morand S, Morse AP, Baylis M. Systematic Assessment of the Climate Sensitivity of Important Human and Domestic Animals Pathogens in Europe. Scientific Reports. 2017;7(1):7134, doi:10.1038/s41598-017-06948-9.

56. Bakhtsiyarava M, Grace K, Nawrotzki RJ. Climate, Birth Weight, and Agricultural Livelihoods in Kenya and Mali. Am J Public Health. 2018;108(S2):S144–s50, doi:10.2105/ajph.2017.304128.

57. Owor BE, Masankwa GN, Mwango LC, Njeru RW, Agoti CN, Nokes DJ. Human metapneumovirus epidemiological and evolutionary patterns in Coastal Kenya, 2007-11. BMC Infect Dis. 2016;16:301, doi:10.1186/s12879-016-1605-0.

58. Wambua J, Munywoki PK, Coletti P, Nyawanda BO, Murunga N, Nokes DJ, Hens N. Drivers of respiratory syncytial virus seasonal epidemics in children under 5 years in Kilifi, coastal Kenya. PLoS One. 2022;17(11):e0278066, doi:10.1371/journal.pone.0278066.

59. Sewe MO, Bunker A, Ingole V, Egondi T, Astrom DO, Hondula DM, et al. Estimated Effect of Temperature on Years of Life Lost: A Retrospective Time-Series Study of Low-, Middle-, and High-Income Regions. Environmental Health Perspectives. 2018;126:17004, doi:10.1289/EHP1745.

60. Egondi T, Kyobutungi C, Rocklöv J. Temperature variation and heat wave and cold spell impacts on years of life lost among the urban poor population of Nairobi, Kenya. Int J Environ Res Public Health. 2015;12(3):2735–48, doi:10.3390/ijerph120302735.

61. Bherwani H, Kumar S, Musugu K, Nair M, Gautam S, Gupta A, et al. Assessment and valuation of health impacts of fine particulate matter during COVID-19 lockdown: a comprehensive study of tropical and sub tropical countries. Environ Sci Pollut Res Int. 2021;28(32):44522–37, doi:10.1007/s11356-021-13813-w.

62. Mukeka JM, Ogutu JO, Kanga E, Piepho H-P, Røskaft E. Long-term trends in elephant mortality and their causes in Kenya. Frontiers in Conservation Science. 2022;3, doi:10.3389/fcosc.2022.975682.

63. Egondi T, Ettarh R, Kyobutungi C, Ng N, Rocklöv J. Exposure to Outdoor Particles (PM2.5) and Associated Child Morbidity and Mortality in Socially Deprived Neighborhoods of Nairobi, Kenya. Atmosphere. 2018;9(9):351, doi:10.3390/atmos9090351.

64. Lee KK, Bing R, Kiang J, Bashir S, Spath N, Stelzle D, et al. Adverse health effects associated with household air pollution: a systematic review, meta-analysis, and burden estimation study. Lancet Glob Health. 2020;8(11):e1427–e34, doi:10.1016/s2214-109x(20)30343-0.

65. Dianati K, Zimmermann N, Milner J, Muindi K, Ezeh A, Chege M, et al. Household air pollution in Nairobi’s slums: A long-term policy evaluation using participatory system dynamics. Sci Total Environ. 2019;660:1108–34, doi:10.1016/j.scitotenv.2018.12.430.

66. Dianati K, Schäfer L, Milner J, Gómez-Sanabria A, Gitau H, Hale J, Langmaack H, Kiesewetter G, Muindi K, Mberu B, Zimmermann N, Michie S, Wilkinson P, Davies M. A system dynamics-based scenario analysis of residential solid waste management in Kisumu, Kenya. Science of the Total Environment. 2021(777):146–200, doi:10.1016/j.scitotenv.2021.146200.

67. Owili PO, Muga MA, Pan WC, Kuo HW. Cooking fuel and risk of under-five mortality in 23 Sub-Saharan African countries: a population-based study. Int J Environ Health Res. 2017;27(3):191–204, doi:10.1080/09603123.2017.1332347.

68. Jiao X, Walelign SZ, Nielsen MR, Smith-Hall C. Protected areas, household environmental incomes and well-being in the Greater Serengeti-Mara Ecosystem. Forest Policy and Economics. 2019;106:101948, doi:10.1016/j.forpol.2019.101948.

69. Mwenda PK, Olago D, Okatcha F, Ali AA. Vulnerability of Communities to Climate Change Induced Disaster Risks and Potential Mental Health Outcomes in Isiolo County, Kenya. Journal of Physical Sciences. 2022;3(1):37–66, doi:10.47941/jps.751.

70. Neale C, Boukhechba M, Cinderby S. Understanding psychophysiological responses to walking in urban settings in Asia and Africa. Journal of Environmental Psychology. 2023;86:101973, doi:10.1016/j.jenvp.2023.101973.

71. Straight B, Hilton CE, Naugle A, Olungah CO, Ngo D, Qiao X, Needham BL. Drought, psychosocial stress, and ecogeographical patterning: Tibial growth and body shape in Samburu (Kenyan) pastoralist children. American Journal of Biological Anthropology. 2022;178(4):574–92, doi:10.1002/ajpa.24529.

72. Egondi T, Kyobutungi C, Kovats S, Muindi K, Ettarh R, Rocklöv J. Time-series analysis of weather and mortality patterns in Nairobi’s informal settlements. Glob Health Action. 2012;5:23–32, doi:10.3402/gha.v5i0.19065.

73. Scott AA, Misiani H, Okoth J, Jordan A, Gohlke J, Ouma G, et al. Temperature and heat in informal settlements in Nairobi. PLoS One. 2017;12(11):e0187300, doi:10.1371/journal.pone.0187300.

74. Asefi-Najafabady S, Vandecar, K. L., Seimon, A., Lawrence, P., & Lawrence, D. Climate change, population, and poverty: Vulnerability and exposure to heat stress in countries bordering the Great Lakes of Africa. Climatic Change. 2018(148):561–73, doi:10.1007/s10584-018-2211-5.

75. Bethancourt HJ, Swanson ZS, Nzunza R, Huanca T, Conde E, Kenney WL, et al. Hydration in relation to water insecurity, heat index, and lactation status in two small-scale populations in hot-humid and hot-arid environments. Am J Hum Biol. 2021;33(1):e23447, doi:10.1002/ajhb.23447.

76. Deribe K, Sultani HM, Okoyo C, Omondi WP, Ngere I, Newport MJ, Cano J. Geostatistical modelling of the distribution, risk and burden of podoconiosis in Kenya. Trans R Soc Trop Med Hyg. 2023;117(2):72–82, doi:10.1093/trstmh/trac092.

77. United States Agency International Development. Climate Risk in Kenya: Country Risk Profile. 2018. Available from: https://www.usaid.gov/climate/country-profiles/kenya.

78. UK Meteorological Office. Climate: Observations, projections and impacts: Kenya. 2011. Available from: https://www.metoffice.gov.uk/binaries/content/assets/metofficegovuk/pdf/research/climate-science/climate-observations-projections-and-impacts/kenya.pdf.

79. Reed SL, McKerrow JH. Why Funding for Neglected Tropical Diseases Should Be a Global Priority. Clinical Infectious Diseases. 2018;67(3):323–6, doi:10.1093/cid/ciy349.

80. Chapman N, Doubell A, Oversteegen L, Chowdhary V, Rugarabamu G, Zanetti R, Ong M, Borri J. Neglected Disease Research and Development: Reflecting on a Decade of Global Investment. Policy Cures Research; 2017. Available from: https://policy-cures-website-assets.s3.amazonaws.com/app/uploads/2020/01/09162349/2017-G-FINDER-report.pdf.

81. Henry J. Kaiser Family Foundation. The U.S. Government and Global Neglected Tropical Disease Efforts. 2020. Available from: https://www.kff.org/global-health-policy/fact-sheet/the-u-s-government-and-global-neglected-tropical-diseases/.

82. Kulkarni MA, Duguay C, Ost K. Charting the evidence for climate change impacts on the global spread of malaria and dengue and adaptive responses: a scoping review of reviews. Globalization and Health. 2022;18(1):1, doi:10.1186/s12992-021-00793-2.

83. Allotey P, Reidpath DD, Pokhrel S. Social sciences research in neglected tropical diseases 1: the ongoing neglect in the neglected tropical diseases. Health Research Policy and Systems. 2010;8(1):32, doi:10.1186/1478-4505-8-32.

84. Charani E, Abimbola S, Pai M, Adeyi O, Mendelson M, Laxminarayan R, Rasheed MA. Funders: The missing link in equitable global health research? PLOS Global Public Health. 2022;2(6):e0000583, doi:10.1371/journal.pgph.0000583.

85. Beran D, Byass P, Gbakima A, Kahn K, Sankoh O, Tollman S, et al. Research capacity building-obligations for global health partners. Lancet Glob Health. 2017;5(6):e567–e8, doi:10.1016/s2214-109x(17)30180-8.

86. O’Neill MS, McMichael AJ, Schwartz J, Wartenberg D. Poverty, environment, and health: the role of environmental epidemiology and environmental epidemiologists. Epidemiology. 2007;18(6):664–8, doi:10.1097/EDE.0b013e3181570ab9.

87. Currie DJ, Smith C, Jagals P. The application of system dynamics modelling to environmental health decision-making and policy - a scoping review. BMC Public Health. 2018;18(1):402, doi:10.1186/s12889-018-5318-8.

88. Hwong AR, Wang M, Khan H, Chagwedera DN, Grzenda A, Doty B, et al. Climate change and mental health research methods, gaps, and priorities: a scoping review. Lancet Planet Health. 2022;6(3):e281–e91, doi:10.1016/s2542-5196(22)00012-2.

89. Government of the Republic of Kenya. Kenya National Adaptation Plan 2015-2030: Enhanced climate resilience towards the attainment of Vision 2030 and beyond. In: Ministry of Environment and Natural Resources, editor. 2016. Available from: https://www4.unfccc.int/sites/NAPC/Documents%20NAP/Kenya_NAP_Final.pdf.

90. Charlson F, Ali S, Benmarhnia T, Pearl M, Massazza A, Augustinavicius J, Scott JG. Climate Change and Mental Health: A Scoping Review. Int J Environ Res Public Health. 2021;18(9), doi:10.3390/ijerph18094486.

91. Bonell AS, Bakary Badjie, Jainaba Samateh, Tida Saidy, Tida Sosseh, Fatou Sallah, Yahya Bajo, Kebba Murray, Kris A Hirst, Jane Vicedo-Cabrera, Ana Prentice, Andrew M Maxwell, Neil S Haines, Andy. Environmental heat stress on maternal physiology and fetal blood flow in pregnant subsistence farmers in The Gambia, west Africa: an observational cohort study. Lancet planetary health. 2022;6(12): e968–e76, doi:10.1016/S2542-5196(22)00242-X.

92. McElroy S, Ilango S, Dimitrova A, Gershunov A, Benmarhnia T. Extreme heat, preterm birth, and stillbirth: A global analysis across 14 lower-middle income countries. Environment International. 2022;158:106902, doi:10.1016/j.envint.2021.106902.

93. Imbo AE, Mbuthia EK, Ngotho DN. Determinants of Neonatal Mortality in Kenya: Evidence from the Kenya Demographic and Health Survey 2014. Int J MCH AIDS. 2021;10(2):287–95, doi:10.21106/ijma.508.

94. United Nations Population Fund. Demographic Dividend: Kenya. The United Nations; 2023. Available from: https://www.unfpa.org/data/demographic-dividend/KE.

95. World Health Organization. Climate change and health research: Current trends, gaps and perspectives for the future. Geneva: World Health Organization (WHO); 2021. Available from: https://www.who.int/publications/m/item/climate-change-and-health-research-current-trends-gaps-and-perspectives-for-the-future.

96. Pearse R. Gender and climate change. WIREs Climate Change. 2017;8(2):e451, doi:10.1002/wcc.451.

97. Harrington LJ, Dean SM, Awatere S, Rosier S, Queen L, Gibson PB, Barnes C, Zachariah M, Philip S, Kew S, Koren G, Pinto I, Grieco M, Vahlberg M, Snigh R, Heinrich D, Thalheimer L, Li S, Stone D, Yang W, Vecchi GA, Frame DJ, Otto FEL. The role of climate change in extreme rainfall associated with Cyclone Gabrielle over Aotearoa New Zealand’s East Coast. World Weather Attribution Initiative Scientific Report. 2023, doi:10.25561/102624.

98. O’Hare R. Somalia drought may have caused more than 20,000 child deaths2023. Available at: https://www.imperial.ac.uk/news/243879/somalia-drought-have-caused-more-than/.

99. Institute Health Metrics Evaluation. Health research by location – Kenya. 2023.Available from: https://www.healthdata.org/research-analysis/health-by-location/profiles/kenya.

100. Schilling J, Werland L. Facing old and new risks in arid environments: The case of pastoral communities in Northern Kenya. PLOS Climate. 2023;2(7):e0000251, doi:10.1371/journal.pclm.0000251.

101. Haider H. Conflict analysis of North Eastern Kenya. Brighton, UK: Institute of Development Studies; 2020. Available from: https://opendocs.ids.ac.uk/opendocs/handle/20.500.12413/15570.

