## Supplemental Table 1 of environmental exposure topic categories and their corresponding subcategories for "Climate-Sensitive Health Outcomes in Kenya: A Scoping Review of Environmental Exposures and Health Outcomes Research, 2000 – 2023"

| **Topic Category** | **Topic Subcategories** |
| --- | --- |
| Climate Change & Seasonality | Climate change |
|  | Seasonality |
|  | Solar radiation |
| Ambient Temperature | Excess heat |
|  | Excess cold |
|  | Temperature variation |
| Rainfall & Humidity | Rainfall |
|  | Humidity |
| Wind Characteristics | Wind speed |
| Flooding & Water Characteristics | Flooding |
|  | Water level change |
|  | Water quality parameters |
| Drought | Drought |
|  | Wildfires |
| Air Pollution | Particulate matter |
|  | NOx, SO2, Ozone |
|  | Cooking fuel emissions |
|  | Lighting emissions |
|  | Second-hand tobacco smoke |
|  | Other air pollutants |
| Terrestrial & Water Pollution | Organic matter contamination |
|  | Inorganic compound contamination |
|  | Microbial contamination |
|  | Plastic pollution |
| Topography | Soil type & moisture |
| Climate Associated Land-use Change | Deforestation |
|  | Urban development |
|  | Habitat & vegetation change |
